# Characteristics of parkrun That Encourage New Participants to Return

**DOI:** 10.1101/2023.03.14.23287282

**Authors:** Andre S. Gilburn

## Abstract

Physical activity is essential to combating the obesity epidemic. parkrun organise free weekly 5km events. Previous studies have identified characteristics of people that act as barriers to new registrants engaging in and returning to events. The current study identifies characteristics of parkrun events that are associated with the return rate of new participants. The return rate of adult first-time participants to parkrun was determined for all events in Scotland over a 1-year period between 2/2019 and 1/2020. A GLMM was used to determine factors associated with whether they returned to parkrun. Return rates were higher for smaller events and events with more other first-time participants. Older participants and male participants were more likely to return. Those that finished in a relatively slow time were disproportionately less likely to return. Events with routes that run alongside freshwater had higher return rates for women. New participants at parkrun are more likely to return if they attended a smaller event suggesting that parkrun should continue to create new events to make parkrun more efficient at retaining new participants. New participants also returned more readily if they attended an event with a high proportion of other adult new participants so specific recruitment events could be advertised to encourage new participants to attend their first event together. New events could be prioritised in proximity to those events that currently experience the highest attendances.

## Introduction

Studies have shown there are more barriers to participating in physical activity for women than men [1,2]. The context in which women will engage in physical activity differs from that of men [3,4]. Motivations for taking part in sport also differ between the genders [5-6]. Encouraging women to participate in sports is important as lack of activity can lead to morbid obesity with 1.4 billion adults estimated to be partaking in insufficient levels of physical activity [7]. Consequently, promotion of wider participation in sport has become a global priority [8].

The environmental context within which people exist can also play an important role in driving the obesity epidemic [9]. These upstream factors can have both negative and positive influences and their management could influence their impacts. For example, the provision of more positive influences, such cycle routes and pleasant spaces in which to exercise, can encourage increased levels of physical activity [9]. Understanding how the genders respond to the provision of positive upstream factors is likely to be crucial to their management and effectiveness [10].

One example of a positive upstream factor is parkrun who organise approximately 2000 weekly 5km events globally [11]. The results of each parkrun event are published on the parkrun website creating a huge dataset on the finishing time, gender and age group of individual participants. In the UK alone when parkrun was suspended in 2020 due to the covid-19 pandemic over 2 million participants had recorded over 30 million runs [12]. Identifying patterns in how individuals respond to their exposure to parkrun could provide important information about how to maximise its effectiveness in promoting physical activity. The parkrun results database is likely to contain potentially important information that could aid better understanding of patterns of participation, for example, identifying the characteristics of those event venues most successful at encouraging first-time parkrun participants to return and attend a second event.

The parkrun dataset contains a huge amount of information on the participation behaviour of the two genders which could be invaluable to identifying barriers to participation and characteristics that encourage new participants to return. A recent study has identified that 43% of people who register for parkrun never take part in an event and a further 22% only participate once [13]. This study questioned those that registered for parkrun to identify barriers to participation and returning [13]. This identified that women, younger adults and the inactive were least likely to participate or return [13].

Studies have revealed that parkrun not only provides an important public health benefit by encouraging physical activity but also improves mental health and well-being 14-18]. Many parkrun routes have sections that run alongside water. Exposure to blue spaces has recently been shown to be particularly effective in improving mental health so routes that run alongside water might provide additional mental health benefits [19].

Previous studies of the parkrun results have generally used descriptive statistics in combination with surveys. The application of more advanced analytical techniques on the parkrun dataset, such as generalised linear modelling, could extract additional potentially important information for both health practitioners prescribing parkrun and to those identifying event venue locations and their routes. These models can quantify the relative importance of different factors and identify interactions between them. They can also be used to partition within and between event venue variance and identify features of events associated with participation [20]. The aim of this study was to use a generalised linear model with a binomial error distribution identify factors associated with first-time participants returning to parkrun.

## Methods

This was an analytical study of aggregated secondary data.

### Data sources

The primary data source was the parkrun results for all events that occurred over a year long period from February 2019 to January 2020 in Scotland [12]. The results for each event were processed using an Excel macro which extracted information about each participant including their age category, parkrun ID number, gender, age group, finishing time, number of participations and date [21]. The Macro extracted the name of participants but this was not harvested or included in the dataset. Adults participating in their first parkrun were identified. Age is provided as a 5-year cohort except for 18-19 year olds. Age was converted to a continuous variable by assigning participants the mid-point for their cohort group. The parkrun results pages provide a link to all participants parkrun history. This was accessed in November and December 2022 meaning all participants had a period of at least 33 months up to a maximum of 46 months after their first participation to return to parkrun. However, it should be noted that Scottish parkrun events were suspended for a period of 17 months between March 2020 and July 2021 because of the SARS-cov2 pandemic. Additional characteristics were collected for the first event each of the participants attended. These were the number of participants, the number of new adult participants, the gender ratio of the participants, the elevation gained on the route, the type of surface and whether a substantial part of the route runs alongside water and whether this is freshwater or saltwater [12]. The dataset consisted of 20,191 adult participants made up of 11,459 females and 8,732 males across 56 different event venues.

### Statistical methodology

The data were analysed using R x64 4.1.1 [22]. A generalised linear mixed model (GLMM) with a binomial error distribution of age graded score was generated using the lme4 function [23]. All continuous explanatory variables were scaled to have a mean of zero and a standard deviation of one including quadratic terms generated for finishing time, number of participants, proportion of first-time participants and date. Event venue was included as a random effect. Minimum Akaike Information Criterion was used to select the optimal model. A general linear model of the finishing time of first-time participants was generated to investigate if this varied with the remoteness of an event and the size of an event using the lm function [24].

## Results

### Factors determining the return rate of first-time parkrun participants

The overall return rate of first-time participants to parkrun was 64.18%. A glm identified a number of significant factors associated with return rate (Table 1). There was a significant increase in return rate with age (Table1, Fig 1). Date was strongly negatively associated with return rate. A significant quadratic term shows that the association with date weakens over time. Male participants (66.5%) were highly significantly more likely to return (Figure 1) than female participants (62.4%). The finishing time at a participants first parkrun was also an important determinant of return rate. The non-significant linear term and significant quadratic term suggests little effect of time on return amongst the faster runners but a disproportionately negative impact of the slowest times on return rates (Figure 2). There was a highly significant quadratic association between the proportion of new participants at an event and return rate, with participants disproportionately more likely to return from events with the highest proportion of new participants (Figure 3). There was also a highly significant association between event size and return rate with first-time participants more likely to return when attending smaller events. The travelling time to the next nearest parkrun also was negatively correlated with return rate. The mean travelling time to the next nearest event was 30 mins for those that returned and 33 for those that did not return. Routes that ran alongside freshwater had significantly higher return rates than events that lacked water and a significant interaction term with gender shows this was particularly the case for women (Table 2). Gender ratio of the field, surface type and elevation gain were not retained in the model.

**Table 1.**
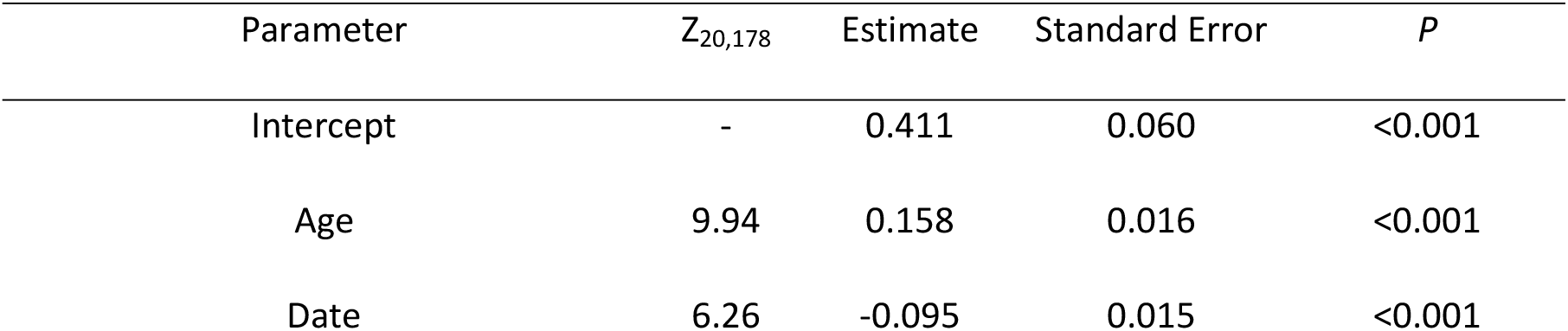
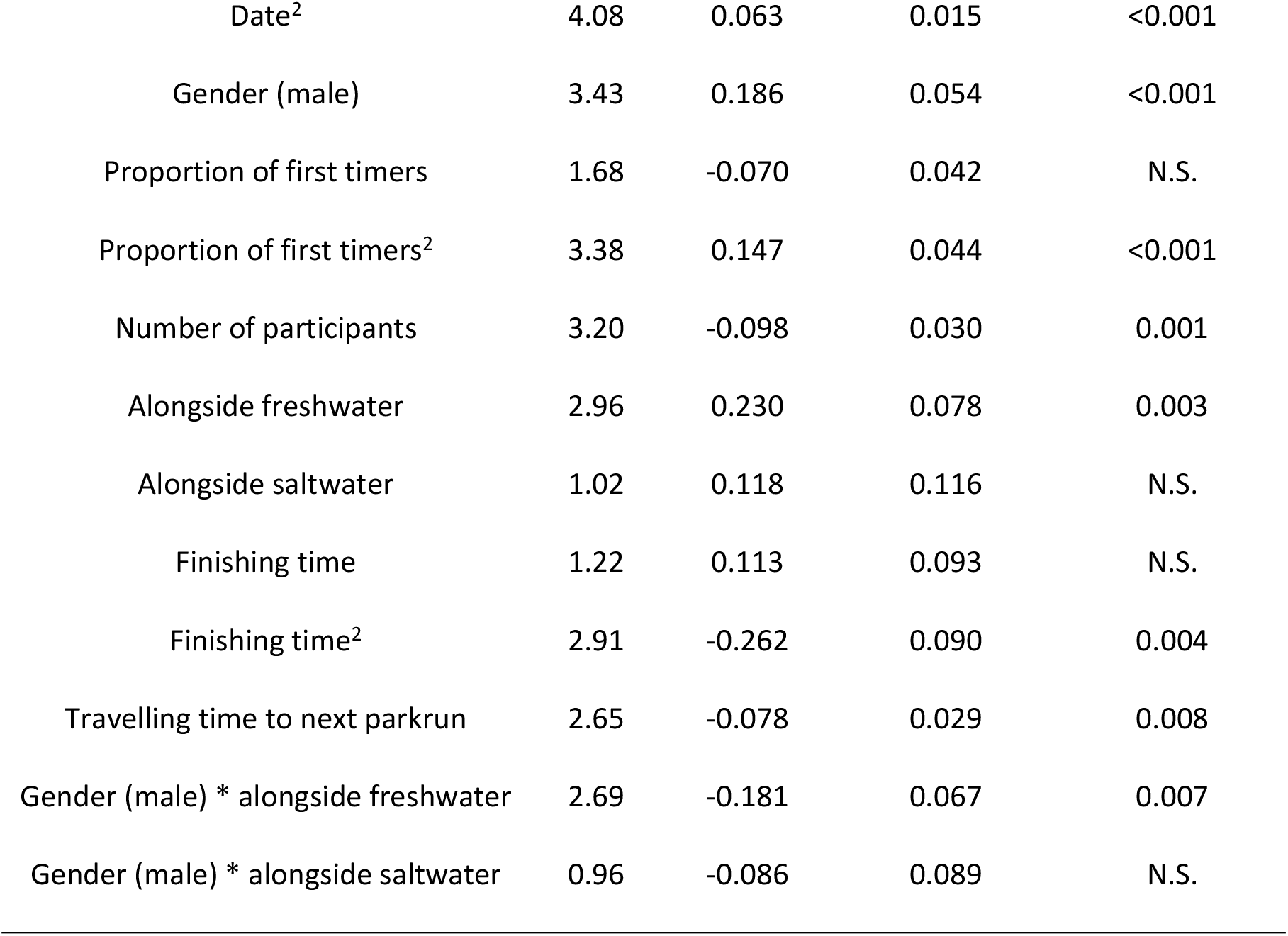
A generalised linear model with binomial error distribution of return rate to parkrun of adult first-time participants. All continuous explanatory variables were scaled. A single interaction term and three quadratic terms were retained in the model.

**Table 2.** The return rate of adult first time parkrunners by gender and route type categorised by whether the route runs alongside freshwater, saltwater or no water.

| Route type | Gender | Return Rate | S.E. | N |
| --- | --- | --- | --- | --- |
| Alongside freshwater | Female | 0.643 | 0.006 | 5812 |
| Alongside saltwater | Female | 0.596 | 0.011 | 1864 |
| Not alongside water | Female | 0.609 | 0.008 | 3783 |
| Alongside freshwater | Male | 0.667 | 0.007 | 4216 |
| Alongside saltwater | Male | 0.637 | 0.012 | 1497 |
| Not alongside water | Male | 0.675 | 0.009 | 3019 |

**Fig 1.**
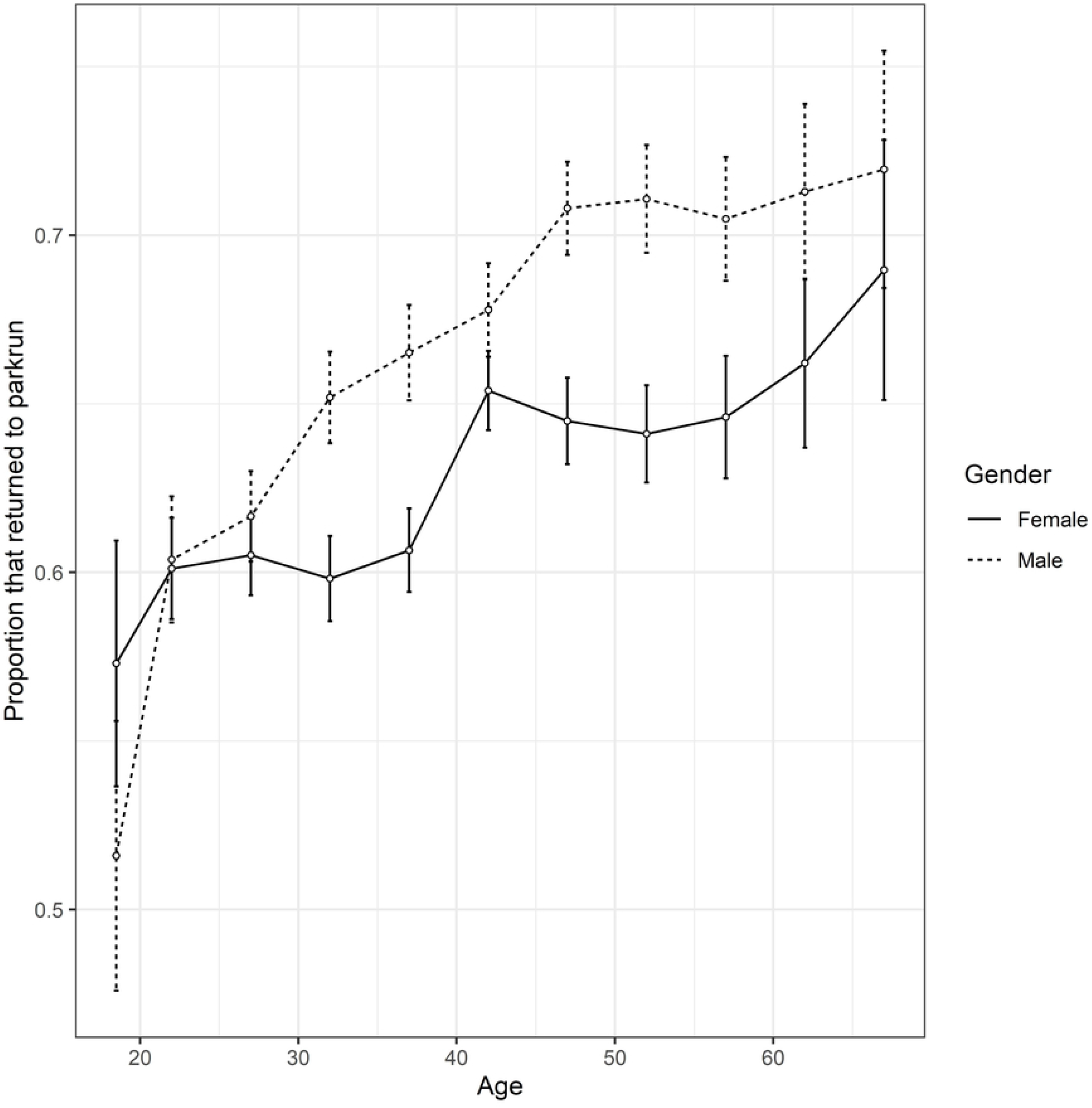
The return rate of adult first-time participants to parkrun events in Scotland against their age. Standard error bars are provided for the mean return rate for each age cohort for both sexes.

**Figure 2.**
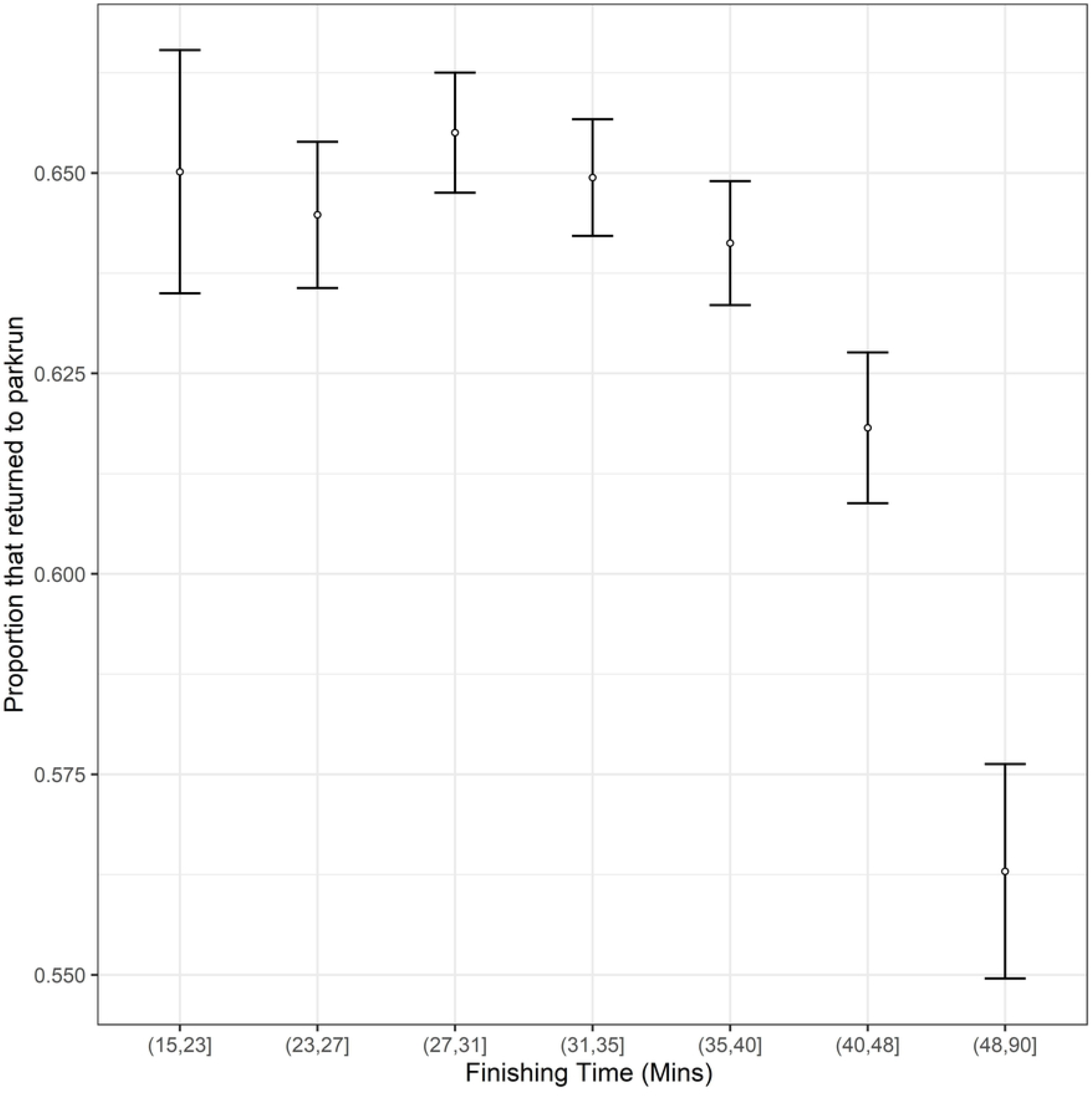
The return rate of adult first-time participants to parkrun against their finishing time. Standard error bars are provided for cohorts based upon finishing time. N.B. Finishing time was treated as a continuous variable in the analyses. Cohorts have simply been created to aid illustration of the data.

**Figure 3.**
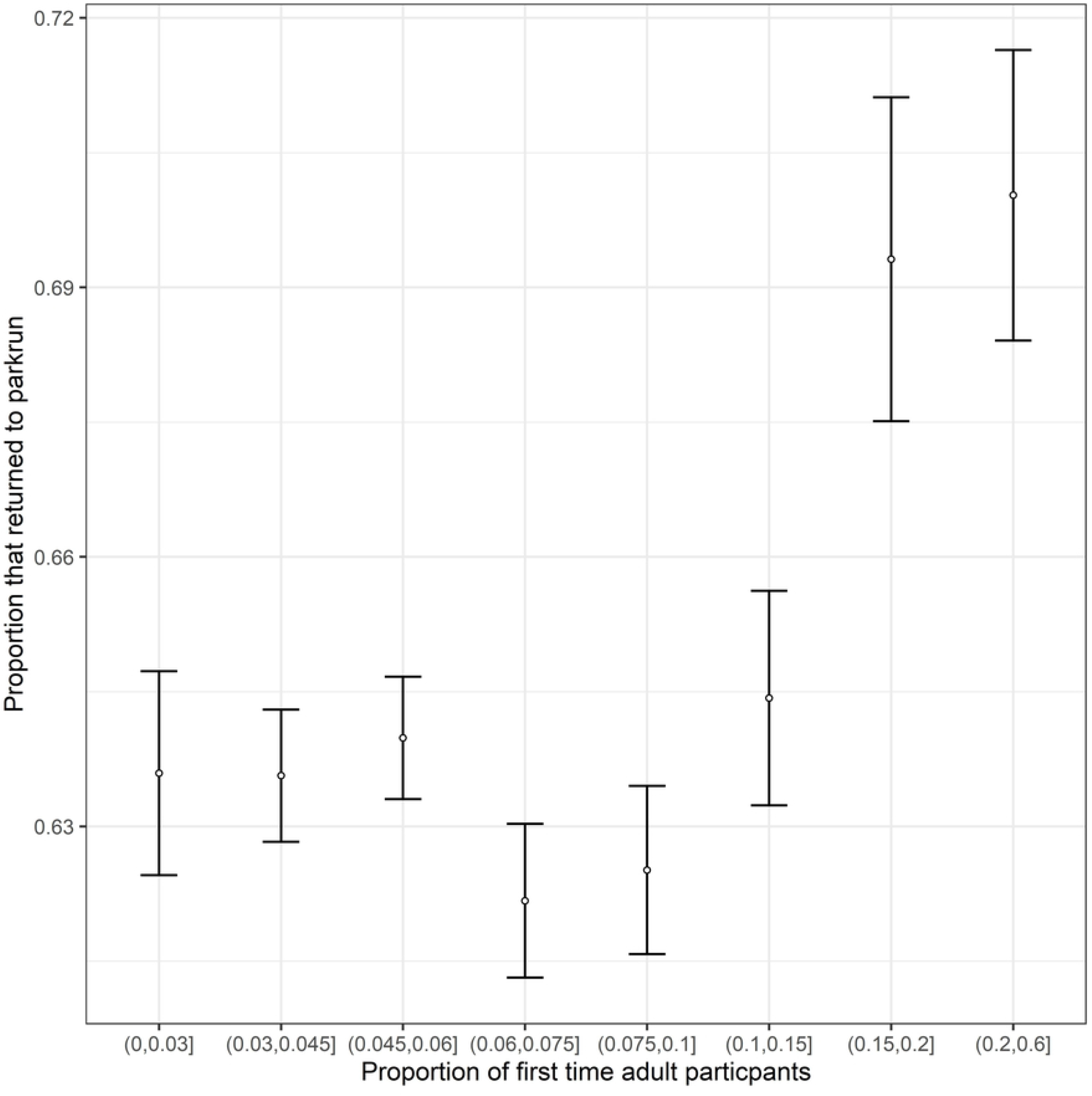
The proportion of first-time adult participants to Scottish events that returned to parkrun against the proportion of new adult participants at the event venues they attended. Standard error bars are provided for cohorts based upon the proportion of adult participants attending events. N.B. Proportion of new adult participants was treated as a continuous variable in the analyses. Cohorts have simply been created to aid illustration of the data.

### Factors Determining Choice of First parkrun Venue

A general linear model was generated that revealed that events with larger number of runners had faster finishing times (F_1,20188_ =231.4, P<0.001) for first time participants and that more remote events (F_1,20188_ =14.9, P<0.001) had slower finishing times for first time participants.

## Discussion

### Return rates to parkrun

The overall return rate of 64.18% is higher than the 61.95% reported in an earlier study [13]. In the current study participants had a longer period of time within which to return (33-46 months compared to 2 years). However, Scottish parkruns were suspended for 17 months during the pandemic meaning that the possible return period was effectively 16-29 months which is reasonable comparable to, albeit on average slightly shorter than, the previous study. This suggests that Scottish parkruns might be relatively effective at getting new participants to return to parkrun. There was an association between date and return rate showing those that had a longer time to return had a slightly higher return rate suggesting that some first timers do eventually return to parkrun a few years after they attended their first event.

### Characteristics of first-time participants associated with return rate to parkrun

Age and gender were both found to affect likelihood of returning to parkrun for first-time participants in Scotland. This is consistent with the finding of a larger scale study covering the UK, Ireland and Australia [13]. The gap in return rate between the genders was slightly wider in Scotland (66.5% for males, 62.4% for females) compared to the broader geographic study that found return rates of 63.7% for men and 60.4% in women [13]. The wider gap in Scotland might be indicative of a larger gender gap in activity in Scotland.

The broader geographic study found from surveying participants that those that engaged more readily in physical activity were more likely to return. The current study found the finishing time of first-time participants were associated with return rate with the return rates of those with finishing times of over 40 minutes (Figure 2) disproportionately low. Those that exercise more regularly are likely to be fitter, and therefore run faster times, so this finding is also consistent with the previous study suggesting that the least active, and those most likely to benefit from parkrun, are those that are least likely to return [13].

### Characteristics of parkrun events associated with the return rate of new participants

This study was the first to investigate what characteristics of parkrun events are associated with the return rate of first-time parkrun participants. Those who attended events with a higher proportion of other adult first-time parkrunners were more likely to return. First time participants who attended an event where the travelling time to the next nearest event was lower were also more likely to return. This suggests that density of parkruns within a local area could be influencing return rates with individuals more likely to return to parkrun in areas with a higher density of event venues. Individuals who enjoyed the general experience of parkrun but maybe not the specific course at the venue they attended could be more likely to try a different parkrun if they have another local alternative.

First-time parkrun participants were also more likely to return after attending events with smaller field sizes suggesting that smaller events might feel more welcoming to new participants. The study also found that first-time participants with the slowest finishing times were more likely to attend smaller and more remote events. This suggests that these events might also appear more attractive to first-time participants as an initial choice of venue as well as providing an experience that is more effective at encouraging them to return.

This study was also the first to try to relate a geospatial feature, whether or not the route ran alongside freshwater or saltwater, to return rate and found that female first-time participants were more likely to return to parkrun if they completed a route with a freshwater blue space. Both exposure to blue spaces and attendance at parkrun have recently been found to be associated with better mental health [15-19]. This study suggests that freshwater blue spaces might well also contribute to the mental health benefit gained by parkrunners by encouraging participants to return.

### Implications for parkrun

These findings suggest that route design could influence whether first timers return. Therefore, parkrun event directors might want to consider these factors when creating new events. The fact that return rates were higher at smaller events and at events where the travelling time to other events was lower both support the continued creation of more parkrun venues. This recommendation does assume that local areas can sustain additional events by finding enough volunteers willing to contribute to running them.

The discovery that new participants are more likely to return after attending an event with a higher proportion of other first-time participants suggests that parkrun might want to consider introducing specific event days when new participants are particularly encouraged to attend so the proportion of new attendees is higher.

Another implication of the findings of this study relates to the parkrun practice initiative^24^. Practitioners utilising the parkrun practice might want to consider prescribing specific local event parkrun events that could particularly increase the patient’s likelihood of returning, for example, by recommending smaller events, attending events with friends also considering parkrun and, in the case of female patients, events that include a section alongside freshwater.

### Limitations of the study

The study was limited to a year-long period due to the considerable time needed to generate the dataset. It is known that the gender gap in participation has been narrowing and this could be partly driven by changes in the return rate of the different genders over time. Therefore, it would be useful to compare this study to other time periods to determine how general the findings are. It is notable in this study that although return rates were lower for female first-time participants there were still more returning female than male first-time participants because the difference in return rate was more than compensated by the higher proportion of female first-time participants. Therefore, the study period was associated with a clear narrowing of the gender gap and shows that, at least in Scotland, the majority of new parkrunners that return to parkrun are actually female which is really encouraging and suggests that parkrun is successful at overcoming traditional barriers to female participation in sports. The restriction of the study to Scotland is another limitation. It would be useful to conduct similar analyses of parkrun return rates in other areas to determine the generality of the findings.

The sars-cov-2 pandemic resulted in parkrun being suspended in Scotland for a period of 17 months. This could have impacted the return rates of participants. The sample duration for first-time participants was specifically chosen to cover the full one-year period before news of the potential pandemic hit the media to make the findings reasonably current but without being the pandemic influencing the sample. Therefore, all participants attended their first event without knowledge of the pandemic, but many will have not returned by the point that news of the emerging pandemic was hitting the media and also at the point that parkrun was suspended in Scotland. The higher return rates found in this study which straddles the covid suspension suggests that it may not have negatively impacted return rates. It would be interesting to investigate the impacts of the pandemic on return rates further by investigating them for first time participants from February 2020 onwards when they would have known about the pandemic.

The single geospatial characteristic of events used in the study was crude in simply considering whether the route ran alongside fresh or saltwater. Therefore, the associations found should be treated with caution. A more detailed geospatial analysis of various aspects of parkrun routes might well provide a lot more information about why certain events have higher return rates than others. Understanding what processes different events adopt to encourage first timers to return would also be worthy of study.

## Conclusion

Previous studies have highlighted how parkrun is a mass participation event that is having a positive impact on the health and wellbeing of its attendees [25-27]. This study has identified various novel features of parkrun events that could be used to make parkrun even more effective and successful by creating more events, smaller events, higher densities of events and by having specific recruitment days events for new participants.

## Data Availability

The study analyzed aggregated secondary data of existing publicly available datasets. The main dataset is owned by parkrun Global and has been accessed as a permitted act but is not mine to share.

## Acknowledgements

The author acknowledges the use of data owned by parkrun Global. The data have been accessed as a permitted act for independent non-commercial research purposes through fair dealing legislation allowing access to publicly available databases. Only a tiny proportion of the parkrun results database was accessed (data from just 56 of more than 2000 events). The author has no official connection to parkrun but is a regular participant and volunteer.

The author did not collect any personal information but used aggregated secondary data. Ethical approval was obtained from the Stirling University General University Ethics Committee.

## References

1. Yungblut HE, Schinke RJ, McGannon KR. Views of adolescent female youth on physical activity during early adolescence. J Sports Sci Med. 2012; 11: 39–50.

2. O’Reilly N, Brunette M, Bradish C. Lifelong Female Engagement in Sport A Framework for Advancing Girls’ and Women’s Participation. J Appl Sport Man. 2018; 10:15–30.

3. Aaltonen S, Waller K, Vähä-Ypyä H, Rinne J, Sievänen H, Silventoinen K, Kaprio J, Kujala UM. Motives for physical activity in older men and women: A twin study using accelerometer-measured physical activity. Scand J Med Sci Sports. 2020; 30: 1409–1422.

4. Burke SM, Carron AV, Eys MA. Physical activity context: Preferences of university students. Psych Sport Exer. 2006; 7: 1–13.

5. Saez I, Solabarrieta J, Rubio I. Motivation for Physical Activity in University Students and Its Relation with Gender, Amount of Activities, and Sport Satisfaction. Sustainability. 2021; 13: 3183.

6. Sattler KM, Deane FP, Tapsell L, Kelly PJ. Gender differences in the relationship of weight-based stigmatisation with motivation to exercise and physical activity in overweight individuals. Health Psych Open. 2018; 5: 2055102918759691

7. Guthold R, Stevens GA, Riley LM, Bull FC. Worldwide trends in insufficient physical activity from 2001 to 2016: a pooled analysis of 358 population-based surveys with 1·9 million participants. Lancet Glob Health. 2018; 6: 1077–86.

8. WHO. Global Action Plan on Physical Activity 2018–2030: More Active People for a Healthier World. Geneva, Switzerland: World Health Organization (WHO); 2018.

9. Lakerveld J, Mackenbach J. The Upstream Determinants of Adult Obesity. Obes Facts. 2017; 10: 216–222.

10. Crawford D, Ball K. Behavioural determinants of the obesity epidemic. Asia Pac J Clin Nutr 2002; 11: S718–S721.

11. parkrunGlobal. https://www.parkrun.com/

12. parkrunUK. https://www.parkrun.org.uk/

13. Recce LJ, Owen K, Graney M, Jackson C, Shields M, Turner G, Wellington C. Barriers to initiating and maintaining participation in parkrun. BMC Public Health. 2022; 22: 83

14. Stevinson C, Hickson M. Parkrun, activity and health: The public health potential of parkrun. J Phys Act Health. 2018; 15: S153

15. Grunseit A, Richards J, Merom D. Running on a high: parkrun and personal wellbeing. BMC Public Health. 2018; 18: 59.

16. Morris P, Scott H. Not just a run in the park: a qualitative exploration of parkrun and mental health. Adv Ment Health. 2019; 17: 110–123.

17. Dunne A, Haake S, Quirk H, Bullas A. Motivation to Improve Mental Wellbeing via Community Physical Activity Initiatives and the Associated Impacts-A Cross-Sectional Survey of UK parkrun Participants. Int J Environ Res Public Health. 2021; 18: 13072.

18. Stevinson C, Hickson M. Changes in physical activity, weight and wellbeing outcomes among attendees of a weekly mass participation event: a prospective 12-month study. J Public Health. 2019; 41: 807–814.

19. McDougall CW, Hanley N, Quilliam RS, Bartie PJ, Robertson T, Griffiths M, Oliver DM. Neighbourhood blue space and mental health: A nationwide ecological study of antidepressant medication prescribed to older adults. Landsc Urban Plan. 2021; 214: 104132

20. Gilburn AS. New parkrunners are slower and the attendance gender gap narrowing making parkrun more inclusive. Int J Environ Res Public Health. (forthcoming).

21. Hoffman R. https://www.facebook.com/groups/parkrunstatsgeeks/files version published October 31st 2019.

22. R Core Team. R: A language and environment for statistical computing. R Foundation for Statistical Computing. Vienna, Austria. URL https://www.R-project.org/. Accessed (2021).

23. Bates D, Mächler M, Bolker B, & Walker S. Fitting Linear Mixed-Effects Models Using lme4. J Stat Softw. 2015; 67: 1–48.

24. https://www.rdocumentation.org/packages/stats/versions/3.6.2/topics/lm

25. Fleming J, Wellington C, Parsons J, Dale J. Collaboration between primary care and a voluntary, community sector organisation: Practical guidance from the parkrun practice initiative. Health Soc Care Community. 2021; DOI: 10.1111/hsc.13236

26. Reece LJ, Quirk H, Wellington C, Haake SJ, Wilson F. Bright spots, physical activity investments that work: parkrun; a global initiative striving for healthier and happier communities. Br J Sports Med. 2019; 53: 326–7.

27. Stevinson C, Hickson M. Exploring the public health potential of a mass community participation event. J Public Health. 2013; 36: 268–274.

28. Grunseit AC, Richards J, Reece L, Bauman A, Merom D. Evidence on the reach and impact of the social physical activity phenomenon parkrun: A scoping review. Prev Med Rep. 2020; 20: 101231

